# Physician burnout in the context of the COVID-19 pandemic: prevalence and associated factors among resident doctors and consultants in Delta State, Nigeria

**DOI:** 10.1101/2024.05.15.24307433

**Authors:** Nnamdi Stephen Moeteke, Ezinneamaka Erhirhie

**Affiliations:** Department of Community Medicine, Delta State University Teaching Hospital, Oghara, Delta State, Nigeria; Department of Family Medicine, Delta State University Teaching Hospital, Oghara, Delta State, Nigeria

**Author notes:** Department of Community and Public Health, Idaho State University, Pocatello, Idaho, United States (Nnamdi Stephen Moeteke). **Corresponding author** (NSM).

**Keywords:** Burnout, Physicians, Delta State, COVID-19

## Abstract

**Background:** Residents Doctors (RDs) and consultants carry out the most specialised medical care. The strain of their job predisposes them to the three domains of burnout: Emotional Exhaustion (EE), Depersonalisation (DP), and diminished Personal Accomplishment (PA). Globally, this public health crisis has worsened with the overwhelming effect of COVID-19 on health systems.

**Aim:** This study assessed the prevalence and associated factors of burnout among RDs and Consultants in tertiary hospitals in Delta State, Nigeria during the pandemic.

**Methods:** A cross-sectional design was employed. Previously validated instruments were used to collect data via an online survey. The questionnaire was sent to physicians selected by multistage sampling. The proportion of participants with a high grade in each of the domains of burnout was obtained. Stepwise analyses from bivariate to multivariate were done to obtain adjusted odds ratios.

**Results:** The prevalence of high-grade burnout in EE, DP, and PA was 35.1%, 13.2%, and 33.3% respectively. Relative to those ≤ 30 years, the age group 41 – 50 years had less likelihood of high EE (AOR 0.050; 95% CI 0.004 – 0.651). Other independent predictors of high EE were manageable workload (AOR 0.094; 95% CI 0.027 – 0.328), reward for work (AOR 0.427; 95% CI 0.205 – 0.892), and good leadership (AOR 0.525; 95% CI 0.113 – 0.929).

**Conclusion and Contribution:** This study suggests that the determinants of burnout among RDs and consultants are mainly contextual factors in the work setting. Promoting an institutional culture of leadership, a manageable workload, and appropriate rewards could help control physician burnout.

## INTRODUCTION

In Nigeria, a resident doctor (RD) is a registered medical/dental graduate working and undergoing postgraduate clinical training (under consultant supervision) in a particular specialty at a health facility accredited by the regulatory bodies for postgraduate medical education.^1^ A consultant is a medical/dental practitioner who, having completed the residency training, works as a specialist in a health facility or medical school.^2^ RDs (registrars and senior registrars) and consultants are involved in the entire spectrum of patient care, but mainly in highly specialised and critical care in tertiary health facilities. There are about 17,000 RDs (making up 40% of the doctor population in the country) and over 4000 consultants in Nigeria.^3,4^

Medical practice and its demands of saving lives can be psychologically stressful.^5^ Long on-the-job hours, including call duties, are the norm.^6^ These strains are amplified in developing countries like Nigeria with constantly unconducive conditions - lack of essential tools/equipment, inefficient systems, poor remuneration, and absence of support, etc. - which lead to incessant brain drain and more workload for the remaining manpower.^6,7^ These predispose physicians and other healthcare professionals to burnout. Burnout is characterised by a combination of associated symptoms emanating from long-standing poorly managed workplace stress, with three facets namely:

a. Feelings of energy depletion/exhaustion (emotional exhaustion, EE)
b. Increased emotional distance from one’s job, or feelings of negativism/cynicism related to one’s job (Depersonalisation, DP)
c. Reduced professional self-esteem (low personal accomplishment, PA).^8^

Globally, burnout in health professionals has become more rampant following the COVID-19 pandemic, with many forced to work in overwhelmed health systems, with high levels of risk, anxiety, and mortality.^9^ The impact of burnout on physicians includes depression and poor general health, individual and workplace impairment, substance abuse, and suicide. These ultimately affect the quality of care of patients (through decreased professional capacity, indifference and loss of empathy for patients, and propensity to healthcare errors), the health organisations and systems (absenteeism and high turnover of already trained personnel), and colleagues and families of affected doctors (relationship friction), making it a ‘public health crisis’.^10–13^ A survey conducted in Eastern Nigeria just before the pandemic got to the country revealed that the prevalence of burnout was 69% among healthcare workers, with as high as 40% screening positively for depression, and the domains of burnout correlating strongly with decreased productivity.^14^ The latest ‘Physicians’ Oath’ acknowledges the welfare of the medical doctor, and inductees are now required to swear to “attend to my own health, well-being, and abilities in order to provide care of the highest standard”.^15^ However, unlike somatic ailments, smouldering symptoms of burnout are often put on the back burner, especially in developing countries where the condition is under-reported.^16^ Though the term “burnout” has been around since the 1970s, it is acknowledged that more data is continually required to design evidence-based control measures among physicians.^5^

The Maslach Burnout Inventory (MBI), a 22-item questionnaire, is the most prominent tool for measuring burnout as defined by the World Health Organisation (WHO) and the gold standard.^13,17^ It has been validated by countless research on burnout done since the tool was developed over three decades ago.^18^ MBI measures burnout in each of the three dimensions using questions with responses on a 7-point scale: ‘Never’, ‘a few times a year’, ‘once a month’, ‘a few times per month’, ‘once a week’, ‘a few times a week’, and ‘every day’, with the scores ranging from 0 - 6. There are nine questions for ‘Emotional Exhaustion’ (EE), five for ‘Depersonalisation’ (DP), and eight for ‘Personal Accomplishment’(PA).^19^ The original MBI Manual categorises scores in each domain into “low,” “moderate,” and “high” burnout (Table 1).^20^ The different dimensions are non-cumulative.

**Table 1:** Categorisation of MBI Scores.

| **Subscale** | **Category** | **Cut-off Scores** |
| --- | --- | --- |
| **EE**<br>**(Score: 0 – 54)** | High | $\geq 27$ |
|  | Moderate | 19 – 26 |
|  | Low | 0 – 18 |
| **DP**<br>**(Score: 0 – 30)** | High | $\geq 10$ |
|  | Moderate | 6 - 9 |
|  | Low | 0 - 5 |
| **PA**<br>**(Score: 0 – 48)** | High | 0 - 33 |
|  | Moderate | 34 - 39 |
| | Low | $\geq 40$ |

The absence of a universal interpretation of the MBI, whereby overall burnout can be presented as a single binary variable, causes problems. While some investigators describe overall burnout as the presence of high burnout in all three domains, others have considered study participants to be truly burnt out if at least two sub-scales are in the ‘high’ category.^21,22^ Other authors see burnout as having either high emotional exhaustion or high depersonalisation.^23^ Overall burnout has also been estimated based on participants who had high burnout in at least one of the three domains.^19^ These variations make it more difficult to make comparisons between studies, with some studies whose authors belong to the first two schools of thought profoundly underestimating rates, at least potentially.^24^ While the developer of the instrument supports the idea of defining clinical burnout as the presence of a high EE burnout alongside either a high DP or PA burnout, she recommends that when assessing for associations between burnout and other variables, it is best to use individual subscale scores as numerical data, or, less preferably, to report burnout in each domain as a categorical variable based on the already established cut-offs for ‘high’, ‘moderate’, and ‘low’.^24^

It is difficult to ascertain the prevalence of overall burnout among physicians in low- and low-middle-income countries due to variations in research instruments and criteria. Nine out of the studies obtained from the literature search employed the MBI and the true prevalence of high EE, DP, and PA burnout can only be estimated from these studies. The papers that examined both RDs and consultants like this current study reported levels of high EE, DP, and PA burnout ranging from 20% - 39.4%, 30% - 71%, and 25% - 50.3% respectively.^25–28^

The relationship between socio-demographic characteristics and physician burnout is unclear as the evidence varies.^19,25,29–31^ However, the higher likelihood of burnout among younger physicians and those with fewer years in practice has been highlighted.^25,26,29,32^ More consistent predictors of physician burnout are work- and training-related factors such as frequency/duration of being on call, number of work hours/workload, remuneration/economic problems, organisational support, recognition from hospital managers, getting adequate skill development and professional training including pandemic-related training, satisfaction with hospital infection control measures, and frequency in dealing with suspected, confirmed, critical cases of COVID-19. These indicate that determinants of burnout are more of contextual factors related to organisational arrangements and management systems in the work setting.^33^ This is corroborated by the qualitative aspect of the study by Ghazanfar et al.^34^ In low- and low-middle-income countries, only five studies done in Egypt, Ethiopia, Pakistan, and Sri Lanka explored, apart from workload, different facets of these institutional predictors, albeit superficially.^25,30,32,34,35^ Furthermore, only two studies done in Egypt provide data on the prevalence and associated factors of physician burnout during the COVID-19 pandemic.^25,32^ There is a paucity of studies in Nigeria involving both residents and consultants, assessing organisational factors, and providing data on physician burnout in the context of the pandemic. This study sought to fill the gap in the literature as regards physician burnout during the COVID-19 pandemic and highlight critical areas of strength/weakness in healthcare organisations to guide prevention/control strategies. Therefore, this study aimed to assess the prevalence of burnout among RDs and Consultants working in tertiary health facilities in Delta State, Nigeria, and determine the associated factors such as socio-demographic characteristics, work setting characteristics, and experiences/perceptions working during the pandemic.

## METHODS AND MATERIALS

### Study Design

A cross-sectional design was used.

### Study Setting

This study was done in the two tertiary health facilities in Delta State, Nigeria. These are government-owned multi-specialty hospitals providing the highest level of specialised and critical healthcare to inhabitants of the state and its environs. These facilities had 502 RDs and Consultants.

### Study Population/Selection Criteria

RDs and consultants who had worked in the facilities for ≥ 1 year were included. Supernumerary RDs and consultants hired on a contractual or part-time basis were excluded.

### Sample Size Calculation and Sampling

For identifying a proportion in a population:

n = 1.96^2^ P(100-P)/E^2^ where

n = minimum sample size

P = proportion of the population with burnout (75.5%) from previous work^31^

E = acceptable margin of error (5%)

n = 1.96^2^ x 0.755 x 0.255/0.05^2^ ≈ 296

Since the total number of RDs/Consultants is less than 10,000, the following correction formula was used to obtain a corrected sample size, n_c_.

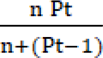

Where n is the uncorrected sample size, and Pt is the total population of RDs/consultants.

n_c_ = (296 × 502)/ (296 + [502 – 1]) ≈ 187

Assuming a non-response rate of 10%, this sample size was increased by 10% to 206.

Based on the total number of RDs and consultants in each hospital, the total number in each specialty, and the numbers of RDs and consultants in each specialty, 206 RDs/consultants were selected by multistage sampling technique. This involved stratified sampling in three stages to proportionately allocate participants to the two hospitals; to individual specialties in each hospital; and, to the three cadres (registrar, senior registrar, and consultant) in each specialty within each hospital.

### Recruitment

The directory of the physicians’ association in the state (to which these researchers belong) which is readily available to members was used. All doctors have access to this document as a shared resource, and these researchers are permitted to send recruitment invites out to the list with no need for a gatekeeper. The study was advertised in the doctors’ WhatsApp groups with a brief description of the purpose, aim/objectives, eligibility criteria, and contact details of the researcher. Thereafter, the web link to the online participant information document (PID), consent form, and questionnaire was sent via WhatsApp to the selected individuals, with a friendly reminder after two, four, and six weeks respectively. Data was collected between 13^th^ January and 5^th^ March 2022.

### Method of Data Collection

Transform™ Survey Hosting was used.^36^ The questionnaire (about 25 minutes to complete) consisted of four sections: socio-demographic/personal professional data, the Maslach Burnout Toolkit™, and the Pandemic Experience and Perceptions Survey (PEPS). Maslach Burnout Toolkit™ “combines the Maslach Burnout Inventory™ (MBI) and the Areas of Worklife Survey (AWS) to measure burnout in the work-life context”. ^37^

The AWS, which has proven validity/reliability in different occupational settings, is a companion tool to the MBI and assesses workplace characteristics that could predict burnout occurrence.^17^ AWS has 28 items (scored on a five-point Likert scale) which assess the perception of ‘Workload’ (5 items), ‘Control’ (4), ‘Reward’ (9), ‘Fairness’ (6), and ‘Organisational Values’ (4). The average score for each sub-component was obtained.

PEPS has 35 items (scored on a five-point Likert scale) that assess work experiences and perceptions during a pandemic with sub-components which include ‘Extent of Impact’ (3 items), ‘Adequacy of Resources’ (5), ‘Risk Perception’ (7), ‘Worklife’ (7), and ‘Leadership’ (10). The total score for each sub-component was obtained.

### Description of the Dependent Variables

The dependent variables were EE, DP and PA. Burnout was indicated by high category i.e., EE score ≥ 27, or DP score ≥ 10, or PA score ≤ 33.^13^ Each participant was allocated to ‘burnout’ or ‘no burn out’ to create a dichotomous dependent variable for each subscale. The prevalence of burnout was measured by the proportion (percentage) of participants who met the cut-off in at least one subscale.

### Data Analysis

The dataset was checked for missing data and there was none for all the listed variables. Chi-squared test was used to determine if there was an association between categorical independent variables (e.g., age group, marital status) with each domain of burnout as a dichotomous variable. For continuous independent variables (e.g., perception of risk score), association with scores for each domain of burnout was tested using Pearson correlation. To identify independent predictors of each domain of burnout, all categorical variables significant in bivariate analyses were introduced into a binary logistic regression model. The level of statistical significance (α) was set at 0.05 for all analyses.

### Ethical Considerations

Ethical clearance was obtained from the Health Research Ethics Committee of Delta State University Teaching Hospital (Approval Number HREC/PAN/2021/037/0426). Permission to use the MBI, AWS, and PEPS was acquired from the patent holders. Participants had to first go through the Participant Information Document (PID) and consent questions after clicking on the link to the online survey. They were informed that they could withdraw from the study without repercussions and were required to provide written informed consent by ticking checkboxes indicating their consent before proceeding to the questionnaire. The PID had the researcher’s telephone number and email address to enable potential participants ask questions if needed. It also had contact details of support groups/organisations that participants could reach out to if the questionnaire caused distress. The online survey did not have any IP links, so it remained anonymous. There were no identifiers on the questionnaire. Data was password-protected.

## RESULTS

Only 114 persons submitted the filled questionnaire, giving a response rate of 55.3%. The average age of respondents was 40 years (Table 2). Of the 114 respondents, 40 (35.1%) had high EE, 15 (13.2%) had high DP, and 38 (33.3%) had high PA burnout. There was high burnout in at least one domain in 46 persons (40.4%) (Table 3).

**Table 2:** Socio-demographic and professional characteristics of respondents.

|  |  | *TOTAL, N = 114* |  |
| --- | --- | --- | --- |
|  |  | *n* | *%* |
| Age | ≤30 years | 10 | 8.8 |
|  | 31 – 40 years | 55 | 48.2 |
|  | 41 – 50 years | 42 | 36.8 |
|  | ≥ 51 years | 7 | 6.1 |
|  | Mean ± SD | 40.30 ± 6.27 |  |
| Sex | Male | 66 | 57.9 |
|  | Female | 48 | 42.1 |
| Marital status | Single | 11 | 9.6 |
|  | Married | 96 | 84.2 |
|  | Separated | 3 | 2.6 |
|  | Divorced | 2 | 1.8 |
|  | Widow | 2 | 1.8 |
| Specialty category | Medical-related | 64 | 56.6 |
|  | Surgical-related | 41 | 36.3 |
|  | Diagnostic | 8 | 7.1 |
| Rank | Consultant | 22 | 19.3 |
|  | Senior resident doctor | 52 | 45.6 |
|  | Junior resident doctor | 40 | 35.1 |
| Number of years post-graduation | ≤10 years | 40 | 35.1 |
|  | 11 – 20 years | 58 | 50.9 |
|  | 21 – 30 years | 15 | 13.2 |
|  | ≥ 31 years | 1 | 0.9 |

**Table 3:** Distribution of burnout categories among respondents.

| Burnout domain |  | Frequency (%) |
| --- | --- | --- |
| **Emotional Exhaustion (EE)** | Low burnout | 53 (46.5%) |
|  | Moderate burnout | 21 (18.4%) |
|  | High burnout | 40 (35.1%) |
| **Depersonalisation (DP)** | Low burnout | 76 (66.7%) |
|  | Moderate burnout | 23 (20.1%) |
|  | High burnout | 15 (13.2%) |
| **Personal Accomplishment (PA)** | Low burnout | 45 (39.5%) |
|  | Moderate burnout | 31 (27.2%) |
|  | High burnout | 38 (33.3%) |
| **Overall Burnout (presence of at least one of High EE, High DP, and High PA burnout)** | Yes | 46 (40.4%) |
|  | No | 68 (59.6%) |

The categorical variables significantly associated with EE were age group and number of years post-graduation (Table 4). Only age group was significantly associated with DP (Table 5). EE scores had a weak positive correlation with the number of work hours per week, and scores for perception of risk; a weak negative correlation with scores for sense of control, community, fairness, values, and adequacy of resources; and a moderate negative correlation with scores for manageability of workload, reward, and experience of leadership. A similar pattern was obtained for DP scores except that all the correlations were weak (Table 6).

**Table 4:** Association between Emotional Exhaustion (EE) and socio-demographic and personal professional characteristics.

|  |  | Emotional Exhaustion |  |  |  |  |  |  |  |
| --- | --- | --- | --- | --- | --- | --- | --- | --- | --- |
| | | | | N = 114 | | | | $\chi^2$ | P-value |
|  |  | Low |  | Moderate |  | High |  |  |  |
|  |  | n | % | n | % | n | % |  |  |
| Age (years) | ≤30 | 2 | 3.8 | 0 | 0.0 | 8 | 20.0 | 18.131 | *0.006 |
|  | 31 – 40 | 23 | 43.4 | 10 | 47.6 | 22 | 55.0 |  |  |
|  | 41 – 50 | 22 | 41.5 | 11 | 52.4 | 9 | 22.5 |  |  |
|  | ≥ 51 | 6 | 11.3 | 0 | 0.0 | 1 | 2.5 |  |  |
| Sex | Male | 27 | 50.9 | 13 | 61.9 | 26 | 65.0 | 2.018 | 0.365 |
|  | Female | 26 | 49.1 | 8 | 38.1 | 14 | 35.0 |  |  |
| Marital status | Single | 4 | 7.5 | 1 | 4.8 | 6 | 15.0 | 5.243 | 0.731 |
|  | Married | 45 | 84.9 | 19 | 90.4 | 32 | 80.0 |  |  |
|  | Separated | 2 | 3.8 | 0 | 0.0 | 1 | 2.5 |  |  |
|  | Divorced | 1 | 1.9 | 1 | 4.8 | 0 | 0.0 |  |  |
|  | Widow | 1 | 1.9 | 0 | 0.0 | 1 | 2.5 |  |  |
| Specialty | Medical | 33 | 63.4 | 10 | 47.7 | 21 | 52.5 | 7.734 | 0.102 |
| *category* | Surgical | 16 | 30.8 | 7 | 33.3 | 18 | 45.0 |  |  |
|  | Diagnostic | 3 | 5.8 | 4 | 19.0 | 1 | 2.5 |  |  |
| *Rank* | Consultant | 14 | 26.4 | 3 | 14.3 | 5 | 12.5 | 8.415 | 0.078 |
|  | Senior Registrars | 24 | 45.3 | 13 | 61.9 | 15 | 37.5 |  |  |
|  | Registrars | 15 | 28.3 | 5 | 23.8 | 20 | 50.0 |  |  |
| *Number of years post-graduation* | ≤10 years | 16 | 30.2 | 4 | 19.0 | 20 | 50.0 | 15.602 | *0.016 |
|  | 11 – 20 years | 24 | 45.3 | 16 | 76.2 | 18 | 45.0 |  |  |
|  | 21 – 30 years | 12 | 22.6 | 1 | 4.8 | 2 | 5.0 |  |  |
|  | ≥ 31 years | 1 | 1.9 | 0 | 0.0 | 0 | 0.0 |  |  |

$\chi^2$ = Chi-squared test statistic | \*statistically significant

**Table 5:** Association between Depersonalisation (DP) and socio-demographic and personal professional characteristics.

|  |  | Depersonalisation |  |  |  |  |  |  |  |
| --- | --- | --- | --- | --- | --- | --- | --- | --- | --- |
|  |  |  |  | N = 114 |  |  |  |  |  |
| | | Low | | Moderate | | High | | $\chi^2$ | P-value |
|  |  | n | % | n | % | n | % |  |  |
| Age (years) | ≤30 | 2 | 2.6 | 3 | 13.0 | 5 | 33.3 | 16.482 | *0.011 |
|  | 31 – 40 | 37 | 48.7 | 12 | 52.2 | 6 | 40.0 |  |  |
|  | 41 – 50 | 32 | 42.1 | 7 | 30.4 | 3 | 20.0 |  |  |
|  | ≥ 51 | 5 | 6.6 | 1 | 4.3 | 1 | 6.7 |  |  |
| Sex | Male | 44 | 57.9 | 14 | 60.9 | 8 | 53.3 | 0.212 | 0.900 |
|  | Female | 32 | 42.1 | 9 | 39.1 | 7 | 46.7 |  |  |
| Marital status | Single | 5 | 6.6 | 3 | 20.0 | 3 | 20.0 | 10.190 | 0.252 |
|  | Married | 67 | 88.2 | 19 | 66.7 | 10 | 66.7 |  |  |
|  | Separated | 2 | 2.6 | 0 | 0.0 | 1 | 6.7 |  |  |
|  | Divorced | 2 | 2.6 | 0 | 0.0 | 0 | 0.0 |  |  |
|  | Widow | 0 | 0.0 | 1 | 4.3 | 1 | 6.7 |  |  |
| *Specialty category* | Medical | 45 | 60.0 | 12 | 52.2 | 7 | 46.7 | 1.277 | 0.865 |
|  | Surgical | 25 | 33.3 | 9 | 39.1 | 7 | 46.7 |  |  |
|  | Diagnostic | 5 | 6.7 | 2 | 8.7 | 1 | 6.6 |  |  |
| *Rank* | Consultant | 16 | 21.1 | 2 | 8.7 | 4 | 26.7 | 8.296 | 0.081 |
|  | Senior Registrars | 39 | 51.3 | 10 | 43.5 | 3 | 20.0 |  |  |
|  | Registrars | 21 | 27.6 | 11 | 47.8 | 8 | 53.3 |  |  |
| *Number of years post-graduation* | ≤10 years | 22 | 28.9 | 9 | 39.2 | 9 | 60.0 | 9.310 | 0.157 |
|  | 11 – 20 years | 42 | 55.3 | 13 | 56.5 | 3 | 20.0 |  |  |
|  | 21 – 30 years | 11 | 14.5 | 1 | 4.3 | 3 | 20.0 |  |  |
|  | ≥ 31 years | 1 | 1.3 | 0 | 0.0 | 0 | 0.0 |  |  |

$\chi^2$ = Chi-squared test statistic | \*statistically significant

**Table 6:** Correlation between burnout scores and continuous independent variables.

|  |  | **Emotional Exhaustion** | **Depersonalization** | **Personal Accomplishment** |
| --- | --- | --- | --- | --- |
| Work hours per week | *r* | 0.231 | 0.186 | 0.057 |
|  | *p-value* | *0.014 | *0.047 | 0.547 |
| Number of calls per month | *r* | 0.017 | 0.153 | -0.101 |
|  | *p-value* | 0.856 | 0.105 | 0.286 |
| Manageability of workload | *r* | -0.538 | -0.269 | -0.129 |
|  | *p-value* | *<0.0001 | *0.004 | 0.173 |
| Control over work life | *r* | -0.309 | -0.278 | -0.023 |
|  | *p-value* | *0.001 | *0.003 | 0.808 |
| Reward for work | *r* | -0.411 | -0.165 | -0.021 |
|  | *p-value* | *<0.0001 | 0.080 | 0.822 |
| Community at workplace | *r* | -0.280 | -0.180 | 0.013 |
|  | *p*-value | *0.003 | 0.055 | 0.890 |
| Fairness at workplace | *r* | -0.326 | -0.225 | 0.070 |
|  | *p*-value | *<0.0001 | *0.016 | 0.459 |
| Workplace values | *r* | -0.307 | -0.252 | 0.008 |
|  | *p*-value | *0.001 | *0.007 | 0.936 |
| Impact of pandemic | *r* | 0.107 | 0.011 | 0.037 |
|  | *p*-value | 0.256 | 0.910 | 0.696 |
| Adequacy of resources | *r* | -0.263 | -0.175 | 0.010 |
|  | *p*-value | *0.005 | 0.063 | 0.912 |
| Perception of risk | *r* | 0.241 | 0.237 | -0.001 |
|  | *p*-value | *0.010 | *0.011 | 0.989 |
| Experience of leadership | *r* | -0.452 | -0.308 | 0.030 |
|  | *p*-value | *<0.0001 | *0.001 | 0.749 |

*r* = Pearson correlation coefficient | \*statistically significant

The multivariate model for high EE showed a decreasing likelihood of high EE with older age groups, though this was statistically significant only for those aged 41-50 years who had a 95% decrease in odds compared to the group of ≤30 years. The AWS and PEPS variables which were independent predictors of high EE were ‘manageability of workload’, ‘reward for work’, and ‘experience of leadership’ (Table 7). There was no statistically significant association between DP and all the predictors entered into the model.

**Table 7:** Binary logistic regression model for factors associated with high Emotional Exhaustion.

|  |  | Adjusted odds ratio | 95 % CI |  | P-value |
| --- | --- | --- | --- | --- | --- |
|  |  |  | Lower | Upper |  |
|  | **Age group** |  |  |  |  |
| **Categorical variables** | ≤30 years* |  |  |  | 0.106 |
|  | 31 – 40 years | 0.189 | 0.021 | 1.688 | 0.136 |
|  | 41 – 50 years | 0.050 | 0.004 | 0.651 | **0.022** |
|  | ≥ 51 years | 0.008 | 0.000 | 1.056 | 0.053 |
|  | **Years post-graduation** |  |  |  |  |
|  | ≤10 years* |  |  |  | 0.784 |
|  | 11 – 20 years | 0.810 | 0.189 | 3.469 | 0.776 |
|  | 21 – 30 years | 3.249 | 0.146 | 72.486 | 0.457 |
|  | ≥ 31 years | 0.000 | 0.000 | . | 1.000 |
| **Continuous variables** | Work hours per week | 1.001 | .986 | 1.017 | 0.862 |
|  | Workload | 0.094 | 0.027 | 0.328 | **0.000** |
|  | Control | 2.118 | 0.838 | 5.352 | 0.113 |
|  | Reward | 0.427 | 0.205 | 0.892 | **0.024** |
|  | Community | 0.799 | 0.343 | 1.861 | 0.602 |
|  | Fairness | 0.843 | 0.266 | 2.674 | 0.772 |
|  | Values | 0.694 | 0.281 | 1.715 | 0.429 |
|  | Adequacy of resources | 1.018 | 0.841 | 1.231 | 0.856 |
|  | Risk perception | 1.047 | 0.936 | 1.170 | 0.424 |
|  | Experience of leadership | 0.525 | 0.113 | 0.929 | **0.043** |

\*Reference category | CI = Confidence Interval

## DISCUSSION

### Implications of the Findings

The findings meet the aims of this study which are to fill the gap in the literature in terms of the prevalence of physician burnout in Nigeria during the pandemic, and, beyond socio-demographic characteristics, to explore predictors such as experiences and perceptions of the pandemic, and workplace characteristics. To the best of our knowledge, this is the first study to provide data in this regard in Nigeria.

The results indicate that the prevalence of high burnout in the EE (35.1%) and PA (33.3%) domains, as well as overall burnout (40.4%), is quite elevated among Nigerian physicians who provide the most specialised and critical care for the populace. However, these levels are slightly less than was obtained in Benin City, Nigeria, and Pakistan,^19,22^ and far less than the other studies in South Africa, Ethiopia, Egypt, and Iran.^28,32,33,35^ The only study with the closest rates (EE 30%, DP 30%, and PA 25%) was that among oncology RDs and consultants in Egypt.^26^ The higher rates in most studies could be because they involved only RDs who are younger doctors and undergo more work stress or focussed on one specialty with a greater tendency for burnout such as emergency medicine.

For the role of socio-demographic factors, there is little discrepancy with other studies that employed multivariate analyses to ascertain independent predictors. Unlike Fernando and Samaranayake,^30^ who observed higher risk among females, and Goyal and colleagues,^38^ who noted a stronger association with the male sex, this study found no gender predisposition in keeping with the majority of other studies.^19,22,25–28,32,33,35^ This study’s observation of significantly higher burnout amongst younger physicians mirrors the findings of others. ^26,27,35^

Unlike in the study by Soltan et al,^26^ where there was no association between burnout and workload, this study showed manageability of workload as a negative correlate of EE, which is in agreement with most others where excessive perceived stress of work overload, higher occupational stress index, dealing with critical cases, more work hours, seeing more patients, and more frequent calls increased burnout.^19,27,28,32,35,38^ Also, the observed negative linear relationship between burnout and reward at work supports other studies that found that burnout has an inverse relationship with recognition from hospital managers, monthly salary,^35^ celebrating accomplishments, having enough money,^22^ and absence of economic problems and concerns about future career.^28^ The absence of an association with control of work life and community is in line with a study that found that burnout is not affected by lack of control over office/training or poor cordiality with colleagues.^22^

Regarding COVID-19 context, though adequacy of resources and perception of risk showed a relationship with burnout on bivariate analysis, these turned out to be insignificant on logistic regression like the study by Mahmood et al.^22^ This is at variance with the finding elsewhere of an association between burnout and receipt of pandemic-related training, satisfaction with organisational infection control precautions, having a colleague who is infected and fear of the coronavirus infection.^25^ The observed protective effect of good leadership is in keeping with other research that highlighted the similar role of a supportive organization and work environment.^22,35^

In general, this study supports the general suggestion in the wider literature that, apart from age, rather than socio-demographic and personal professional variables (such as specialty and years of practice), the determinants of burnout are more of contextual factors related to organizational arrangements and management systems in the work setting. ^33^

### Limitations/Strengths of the Study and Discussion of the Research Process

The strengths of this study include the use of renowned and previously validated instruments which assessed several potential predictors, and the multivariate analyses to ascertain their independent association with the outcomes of interest. Only cross-sectional studies were included in the literature review for direct comparability.

None of the significant associations with DP observed on bivariate analysis was maintained after logistic regression. This may have been due to depreciation in degrees of freedom (and, hence, precision) because the variables included in the regression model were too many for the dataset. It has been suggested that the ratio of events to parameters in a regression model should be 10:1. ^39^ Therefore, with only 15 persons having high DP, having seven variables in the model was likely a pitfall. Another reason for the initially significant predictors becoming insignificant may be due to having two or more input variables that correlate as is most likely the case for age and number of years post-graduation. The consequence is that the effect of each input variable gets dispersed and attenuated.^39^ It would have been better to use only one of the interrelated parameters.

A serious limitation of this study is the low response rate with its implications for the external validity since the sample may be unrepresentative of the study population as individuals who turn down research invitations tend to be different from those who participate.^40^ The extent of this non-response bias could have been determined by comparing the characteristics of responders with the same characteristics of non-responders,^41^ but tangible information about the population was unavailable. The sample size was also below the calculated required minimum which means that it had a reduced statistical power to detect statistically significant differences where such differences exist, and the survey estimates lost precision.^42^ Survey response rates have been on the decline in recent times due to progressively increasing time pressures on people, and medical doctors respond less than other health workers.^42^ This may have affected this study, especially with the shortage of medical personnel due to the massive brain drain and the impact of the COVID-19 pandemic on health personnel/systems. Making repeated contact with selected study subjects is a strategy for improving recruitment,^40^ and this was employed in this study with three reminder WhatsApp messages (at roughly two-week intervals) after the initial contact. These were done courteously, always bearing in mind respect for autonomy.^41^ One person replied that he could not continue with the questions because he found them emotionally discomforting. This suggests that the obtained prevalence of burnout is less than the true picture since non-response, even as low as 25%, distorts estimates of disease burden when the ailment itself is responsible for non-response.^40^ As a duty of care, the physician who experienced emotional distress was directed to organizations he could get help if he thought it necessary. Many of the doctors gave strong feedback that the questionnaire was too long. Indeed, the questionnaire had four sections with a total of 95 questions which, added to the detailed PID and consent form, are cumbersome on a hand-held device. Lengthy questionnaires and online surveys are known to receive lower responses.^42^ More practicably, a shortened form of the MBI could have been used since abbreviated versions have been found to correlate strongly with their respective domain scores from the full MBI and give identical results in the estimation of associations.^13^

### Recommendations

Addressing contextual factors related to organizational arrangements and management systems in the work setting, most importantly workload, leadership, and reward, is crucial for controlling burnout among RDs and consultants in Nigeria. In this regard, the following measures should be adopted by health system managers.

1. More emphasis should be placed on the mental health of post-graduate medical trainees and trainers. There should be an intervention to create more awareness of burnout among RDs and consultants through continuous training programmes that incorporate coping strategies for overcoming work stress and preventing burnout. These should include time management, handling of difficult clinical cases, satisfying home-work demands, mindfulness techniques, cognitive behavioural therapy, etc. Periodic screening as well as counselling systems should also be instituted.^19,29,30,32^
2. Strategies for stress reduction and revision of workload distribution in tertiary hospitals are needed, e.g., limit on number of work hours per week, number of overnight calls per month, etc., and stipulate adequate breaks and vacations.^19,38,43^ Health systems should be structured to make workload manageable by not placing excessively high demands on RDs, or stifling their autonomy at work.^33,44^
3. There is need for a paradigm shift from the traditional paternalistic form of medical leadership which is based only on the merits of academic excellence and professional competence with little or no emphasis on leadership capacity.^45^ With the increasing complexity in medical practice and health systems, medical leadership in Nigeria should also evolve. Appointees to the positions of Chief Medical Director, Director of Clinical Services and Training, Residency Programme Coordinator and other management positions in health care organisations should be persons with vision and well-schooled in critical components of leadership including effective communication, engendering teamwork/supportive organisational culture, and giving adequate recognition to the accomplishments of subordinates.^45,46^ “Professional staff not suited to leadership, either through training, selection or natural inclination should not be entrusted with administrative and management burdens” if possible.^47^ The curricula for medical education, starting from the undergraduate level, should be revised to incorporate training in medical leadership to prepare future managers and enshrine the culture of leadership from the early stages of the profession.^45^
4. Mentorship programmes should be incorporated into the residency training in all accredited health facilities, geared at giving younger doctors all the support they can get since they are more prone to burnout.^48^
5. Adequate reward systems should be designed. Mechanisms should be instituted for the motivation of medical personnel to stimulate job enthusiasm so that they perform their duties with optimum interest and satisfaction in pursuit of organisational and personal goals.^49^ Extrinsic motivation in the form of tangible and intangible incentives from managers of health systems and organisations such as bonuses, recognitions, and salary increases can help control burnout among RDs and consultants in Nigeria, especially against the backdrop of currently poor work conditions that encourage constant brain drain.^1,50^
6. More research on the issue of burnout among RDs and consultants in Nigeria is recommended. Qualitative studies to give a better understanding of these physicians’ experiences of burnout, and longitudinal studies to identify causal factors will support control efforts.

## CONCLUSION

This study examined the prevalence of, and factors associated with burnout among RDs and consultants in the two tertiary hospitals in Delta State, Nigeria. It showed an overall high burnout of 40.4%, with significant predictors being age, workload, leadership, and reward. It suggests significant levels of burnout among a key component of the medical workforce in Nigeria whose mental health and productivity have grave implications for health care. It also reveals predictors that highlight critical areas for intervention in Nigerian healthcare organizations, especially those involved in residency training. Given the dire consequences of physician burnout on patient care, enhancing the work life of RDs and consultants through a framework for stress reduction, amendments in workload distribution, effective leadership, and adequate rewards should be viewed as fundamental to making the best of health system performance, optimizing patient satisfaction, improving population health, and cutting healthcare costs.^22,43,46^

## Data Availability

Will be provided as before publication

## Acknowledgment

This work was part of the research done by Nnamdi Moeteke for his dissertation for the award of the Master of Public Health degree by the University of Liverpool. Special thanks to Dr. Caryl Beynon for the guidance as the Dissertation Advisor!

